# Ambient temperature extremes and neonatal mortality: A time-stratified case-crossover analysis using Demographic and Health Survey data from East Africa (2011-2022)

**DOI:** 10.1101/2025.07.01.25330621

**Authors:** Paerie ul Lokubal, Chérie Part, Chloe Brimicombe, Sari Kovats, Debra Jackson, Hannah Blencowe, Veronique Filippi

## Abstract

**Introduction:** Due to their underdeveloped thermoregulatory system, neonates are at increased risk of morbidity and mortality from hot and cold temperatures. Our study aimed to analyse the effects of environmental temperature on overall, very early, early and late neonatal acute mortality in 5 East African countries using the Demographic and Health Surveys (DHS) data.

**Methods:** We obtained neonatal mortality data from the DHS conducted between 2016 and 2022. Our outcomes were (i) overall neonatal mortality (days 0–27), (ii) very early (day 0); (iii) early (days 1–6); and (iv) late neonatal mortality (days 7–27). Daily mean temperature was constructed from ERA5-Land and assigned at household-level. We used a time-stratified case-crossover design with distributed lag non-linear models (0–6-day lag) to estimate odds of mortality with exposure to the 5^th^ and 95^th^ temperature percentiles (versus the median). Country-level estimates were generated and then pooled to assess the overall association.

**Results:** A total of 1373 neonatal deaths were included, over 80% of which occurred within the first 6 days. The association between ambient temperature and neonatal mortality was heterogenous. In pooled analysis, the 95^th^ and 5^th^ percentiles were associated with increased and decreased mortality odds, respectively, although estimates were imprecise. In Uganda, there was strong evidence of association between high ambient temperature (95^th^ percentile) and overall neonatal mortality (OR=3.54; 95% CI: 1.73–7.28) as well as early neonatal mortality (OR=3.75; 1.70–8.28), while odds of very early neonatal mortality increased with exposure to low temperatures (5th percentile) (OR=5.65; 1.89–16.69). There was no strong evidence of association in other countries.

**Conclusion:** Temperature-related neonatal mortality risk differs across East African countries. Other factors may play a significant role. Future research should consider the effects of environmental temperature on neonatal mortality across different climate zones.

**What is already known on this topic:**

- Hot and cold indoor environments are associated with a risk of neonatal mortality
- Neonatal vulnerability to temperature extremes is most pronounced in the early neonatal period (days 0–6) compared with the late neonatal period (days 7–27)

**What this study adds:**

- This study adds to the body of evidence on the effect of heat on neonatal mortality in different climate regions, allowing for between-country assessment of the association between temperature and neonatal mortality in East Africa.
- Our analysis provides country-level estimates for the association between temperature and overall neonatal mortality, as well as the three distinct phases of the neonatal period (very early, early and late) with differing mortality risk from temperature

**How this study might affect research, practice or policy:**

- There is a need to incorporate heat mitigation strategies into the care for newborn policies and practices, especially for the very early and early neonatal periods

## Introduction

Childhood mortality remains an important global health challenge. The neonatal period, defined as the first 28 days of postnatal life, represents a critical phase for child survival, particularly in low– and middle-income countries (LMICs)[1, 2]. While significant progress has been made in reducing childhood mortality since 1990, the decline in neonatal mortality has been slower than under-5 (U5) mortality[1, 2]. For example, between 2000 and 2023, the global U5 mortality rate decreased by 52% while the neonatal mortality rate (NMR) decreased by 44%[2].

Nearly 50% of all U5 deaths in 2023 were in the neonatal period[2]. In 2023 alone, approximately 2.3 (2.1–2.6) million newborns died within the neonatal period, with the majority occurring in LMICs[1]. Sub-Saharan Africa (SSA) is the world’s region with the highest NMR of 27 deaths per 1000 live births in 2023, compared with 17 deaths per 1000 live births globally and 2 deaths per 1000 live births in Europe and Australia and New Zealand[2]. Within East Africa, NMR remains high and varies between 18 to 22 deaths per 1000 live births, with Kenya and Tanzania having the highest rates at 21.5 (17.4–26.4) and 20.6 (15.6–27.0) respectively, followed by Burundi (19.6, 11.3–34), Rwanda (18.1, 13.5– 24.1) and Uganda having the lowest rate (17.9, 11.2–27.8)[2]. Moreover, at the current pace of decrease in NMR at 34%, the whole of SSA, except South Africa, is at risk of missing the SDG NMR target of ≤12/1,000 live births by 2030[2]. The early timing of most newborn deaths implies a narrow window for life-saving interventions, with about one-third occurring on the day of birth and three-quarters within the first week[1, 3].

The leading medical causes for neonatal mortality include prematurity, birth complications (birth asphyxia), congenital anomalies, lower respiratory infections (pneumonia), and sepsis which account for 86% of all the neonatal mortalities worldwide[1, 2, 4]. Recent research has also demonstrated the significant impact of environmental factors, such as air pollution and environmental temperature, on neonatal mortality[5–8]. Heat extremes are becoming more frequent in Africa[9, 10]. A study bringing together 29 country datasets found that over an 18-year period (2001–2019), 4.3% of neonatal deaths were associated with non-optimal temperatures, with heat and cold responsible for 1.5% and 2.9% of all neonatal deaths, respectively[5]. However, this study used a pooled country data analysis, obscuring between– and within-country temperature differences and its effects on neonatal mortality. Nevertheless, these findings highlight the important role of non-optimal temperatures on neonatal mortality, given that hypothermia) remains high and variable among newborns, ranging from 32–85% among those born in hospitals and 11–92% among those born at home, even in tropical climates [11]. A Norwegian observational study exploring the normal temperature range and risk factors for deviating body temperature during the first 24 hours of life in term-born infants under standardised care showed that, even when delivery room and rooming-in temperatures were maintained at the optimal 26–30°C and 24°C respectively, 28% of infants still experienced hypothermia[12]; highlighting the narrow ambient temperature range needed to maintain neonatal core body temperature at an optimal level.

Neonates are susceptible to environmental temperatures due to their underdeveloped thermoregulatory systems[13, 14]. Their high surface area to volume ratio facilitates rapid heat loss or gain, while limited subcutaneous fat, especially in preterm infants, reduces their ability to maintain core body temperature[3, 13]. The underdeveloped thermoregulatory centre in the hypothalamus, inefficient shivering mechanism, and underdeveloped sweat glands further compromise their ability to regulate body temperature [3, 15]. This vulnerability is most pronounced in the very early (0 – <24 hours postpartum) and early neonatal (1–6 days postpartum) periods when newborns are adapting to the extrauterine environment[13]. There is also evidence that daily temperature extremes exacerbate several causes for neonatal mortality, including respiratory distress and exposure to environmental pathogens, as well as increasing susceptibility to infections, further contributing to neonatal mortality[15, 16]. This risk is even greater for preterm infants and those with low birth weight, who face greater challenges in temperature regulation [3, 12, 17]. In LMICs, additional factors such as inadequate healthcare facilities, lack of temperature-controlled environments, and socioeconomic constraints further exacerbate these risks[5, 18].

Despite the growing recognition of the impact of ambient temperature on neonatal health, there remains a significant gap in understanding how these effects manifest across the very early, early, and late neonatal periods. First, these periods present distinct health challenges to the newborn as explained above[1, 3]. To our knowledge, only one study has explored temperature’s differential impact on neonatal mortality, but it examined only the overall neonatal and very early neonatal periods[5], omitting the distinct early and late neonatal periods. Second, the few existing cross-country studies assessing the effects of temperature on neonatal health outcomes have typically pooled data across countries, which does not allow for between-country differences in the exposure-response relationship[5, 19]. SSA is a large continent with different climate zones, therefore pooling results across them could obscure important regional differences. For example, compared with West Africa, there is less evidence of heat waves in East Africa, although some countries in the region[20], including Uganda and Kenya, are projected to experience some of the greatest increases in the number of hot days[21]. The five countries included in this study experience intra-annual temperature ranges from 11.6°C to 30.7°C, with lower temperatures at high altitudes and higher temperatures in lowland and coastal regions[22]. Kenya and Tanzania, which lie along the coast, have the more diverse temperature ranges, from 10°C to 34°C and 10°C to 32°C, respectively. Uganda averages 17–25°C, while Rwanda and Burundi have milder temperatures, ranging between 10–27°C and 16–23°C, respectively[22]. Our study set out to: (i) explore the impact of ambient temperature extremes on overall, very early, early and late neonatal mortality at country-level; and (ii) compare the effects of ambient temperature extremes on the pooled overall, very early, early and late neonatal mortality in five East African countries using the latest Demographic and Health Survey (DHS) data.

## Methods

### Study population

The DHS are nationally representative household surveys conducted in five overlapping years in over 90 LMICs since 1984[23]. They provide important maternal, newborn and child health information. We conducted a secondary analysis of DHS datasets from five East African countries (Uganda, Kenya, Tanzania, Rwanda and Burundi), conducted between 2016 and 2022[23–25]. We chose the most recent DHS because they include the reported date of birth and date of death for newborns dying within the first 28 days of postnatal life[23–25], which are needed to accurately assign exposures. Women aged 15–49 years are interviewed about their complete birth and child health histories for all the births in the 5 years preceding the survey. Sampling follows a stratified two-stage cluster design: the first stage involved enumeration areas (EAs), also known as primary sampling units (PSUs), which are usually drawn from national census files using probability proportional to size and the second stage involves random sampling of households listed under each EA[25]. The DHS samples are stratified by geographic region and by urban/rural areas within each region. Since early 2000s, the DHS also provides global positioning data (GPS) for each PSU[23]. A PSU is defined as a city block in an urban area and a village in a rural area. To anonymise the respondents, GPS coordinates are randomly displaced by the DHS Program Team. Urban areas are randomly displaced up to 2km and rural areas are randomly displaced up to 5km, with a further 1% displaced up to 10 km[23, 26]. Further details about the DHS survey can be found elsewhere[23–25, 27].

For this analysis, we included all births and deaths occurring within the first 28 days of postnatal life for births within the 5 years preceding the survey in each included country[28].

### Meteorological data

We obtained ERA5-Land 2-metre air temperature and 2-metre dewpoint temperature from the Copernicus Climate Change Service (C3S) of the European Commission, the European Centre for Medium-Range Weather Forecasts (ECMWF) [29]. ERA5-Land is a global gridded reanalysis dataset with a consistently high spatial (0.1° x 0.1°) and temporal (one-hourly) resolution[29]. It is considered highly reliable for capturing temperature trends and variations, demonstrating strong correlation coefficients (0.978 – 0.998) with observed meteorological data[30, 31].

### Exposure and outcomes

Daily mean temperature was constructed from hourly temperature values and used as the main exposure. We adjusted for dewpoint depression as a measure of humidity, defined as the difference between daily mean temperature and daily mean dewpoint temperature. Our outcomes variables were (i) neonatal mortality, (defined as death occurring from day 0 to 27); (ii) very early neonatal mortality, (defined as death occurring on day 0 only); (iii) early neonatal mortality, (defined as death occurring from day 1 to 6); (iv) and late neonatal mortality, (defined as death occurring from day 7 to day 27).

### Study design

We employed a time-stratified case-crossover design, which is used widely to assess acute health effects related to short-term environmental exposures, such as temperature or air pollution [32–37]. The design compares each case’s exposure during the event period to their exposure during control periods when the health event did not occur [38]. Control days were defined as the same day-of-week within the same month and year, resulting in 3-4 control days per case[41]. Thus, by design, this approach adjusts for potential confounding by season and of day-of-week[39–43] as well as individual-level confounders, such as maternal age, education, socioeconomic status, healthcare and other sociodemographic characteristics that are unlikely to change within the small-time windows[33, 34, 40].

### Statistical analysis

DHS and GPS data were linked using the PSU ID. These data were then geospatially linked with the exposure data using the GPS coordinates and date of neonatal mortality. We only linked data for which all GPS coordinates were available, excluding 165 deaths without linkable GPS coordinates.

We used conditional logistic regression, with the PSU and DHS Strata as the ID stratum, to obtain the odds ratio of neonatal mortality at different temperature percentiles compared with the median temperature. We included the DHS strata in our analysis to account for PSU displacement of the GPS coordinates and lack of GPS data on the place of death. Because the DHS is stratified by geographic regions and by urban/rural within each region, we estimated that any mortality, whether at home or hospital, was most likely to occur within the strata or region. We modelled the nonlinear and lagged associations between temperature and neonatal mortality using distributed lag nonlinear models (DLNM) using natural cubic splines and linear terms[44, 45]. For each country, we fitted several models with linear and spline terms (varying degree of freedom (df) and knot placements) in the exposure and lag dimensions. We used the lowest AIC value and examined residuals to determine the best fitting models. For all countries, the best fitting model used a nonlinear term for temperature and a natural cubic spline for the lagged effects with 2 df. All models were adjusted for dewpoint depression. We did not adjust for air pollution due to lack of reliable data in our study settings. Country-level estimates were then pooled using fixed effects to assess the overall association with temperature across all countries.

Finally, the analyses were stratified by wealth group (low-, middle-, and high-income) to assess whether the effect of ambient temperature varied by socioeconomic status. The wealth groups were generated from the DHS quintile groups, with the low-income group generated from the poorest and poor quintiles, middle-income group from the middle quintile, and the high-income group from the richest and richer quintiles.

## Ethics approval

We obtained ethics approval for this analysis from the London School of Hygiene and Tropical Medicine Ethics Approval Committee. LSHTM Ethics Ref: 31865.

## Patient and public involvement

This study had no public or patient involvement, and all the data used in the analysis is anonymised secondary data.

## Results

There were a total of 1538 neonatal deaths in the 5 East African countries recorded in the DHS data sets between 2011 and 2022, with 617 (40.1%) very early, 628 (40.8%) early and 293 (19.1%) late neonatal deaths. After removing cases with missing PSU and GPS data, the total number of neonatal deaths was 1373.

**Table 1:** Summary statistics of neonatal mortality and mean daily temperature.

| Country | Total | Early Neonatal deaths |  |  |  | Late neonatal deaths |  | Temperature (°C) |  |  |
| --- | --- | --- | --- | --- | --- | --- | --- | --- | --- | --- |
|  |  | Very early (Day 0) |  | Early (Day 1-6) |  | Late (Day 7-27) |  | 5<sup>th</sup> per-centile | Median | 95<sup>th</sup> per-centile |
|  | No. | No. | % | No. | % | No. | % |  |  |  |
| Burundi | 279 | 76 | 27.2 | 125 | 44.8 | 78 | 28.0 | 16.7 | 20.6 | 25.0 |
| Kenya | 437 | 179 | 41.0 | 184 | 42.1 | 74 | 16.9 | 17.2 | 25.2 | 30.4 |
| Rwanda | 153 | 81 | 52.9 | 42 | 27.5 | 30 | 19.6 | 16.1 | 19.8 | 22.8 |
| Tanzania | 261 | 97 | 37.2 | 117 | 44.8 | 47 | 18.0 | 17.7 | 22.6 | 27.2 |
| Uganda | 408 | 184 | 45.1 | 160 | 39.2 | 64 | 15.7 | 17.1 | 22.0 | 26.8 |
| Total | 1538 | 617 | 40.1 | 628 | 40.8 | 293 | 19.1 | 17.6 | 22.9 | 28.5 |

No. = Number; % Percentage

As shown in **Table 1**, over 80% of all neonatal deaths occurred within the first week of life; 40.1% of which occurred on day 0. In Rwanda, over 50% of all neonatal deaths occurred within the first 24 hours after birth. Over the study period, the median temperature was 22.9°C (interquartile range (IQR) 20.8–25.2°C) with a 5th percentile of 17.6°C and a 95th percentile temperature of 28.5°C. Kenya had the highest median temperature during the surveys at 25.2°C (IQR 21.7–27.8°C) followed by Tanzania and Uganda at 22.6°C (20.7– 24.5°C) and 22.0°C (20.4–23.6°C), respectively. Rwanda had the lowest median temperature at 19.8°C (18.1–21.2°C) followed by Burundi at 20.6°C (19.1–22.3°C).

Additional details shown in **ST 1** in the supplementary material.

Figure 1 shows the association between daily mean temperature exposures 0-6 days before death and the odds of neonatal mortality (0-27 days postpartum), centred at the median temperature. Overall, we observed between-country heterogeneity in the exposure-response association. In Uganda, lower temperatures were associated with decreased odds of neonatal mortality, while higher temperatures were associated with increased odds (Figure 1a). There was no strong evidence of association in other countries, although there was a tendency towards heterogenous effects. Tanzania and Rwanda (Figures 1b and 1c) showed a tendency towards increased odds of mortality with increasing temperatures, while we observed the opposite in Burundi and Kenya (Figures 1d and 1e). Exposure-response curves for very early, early and late neonatal mortality are provided in the supplementary materials **(ST 2 – ST 4).**

**Figure 1:**
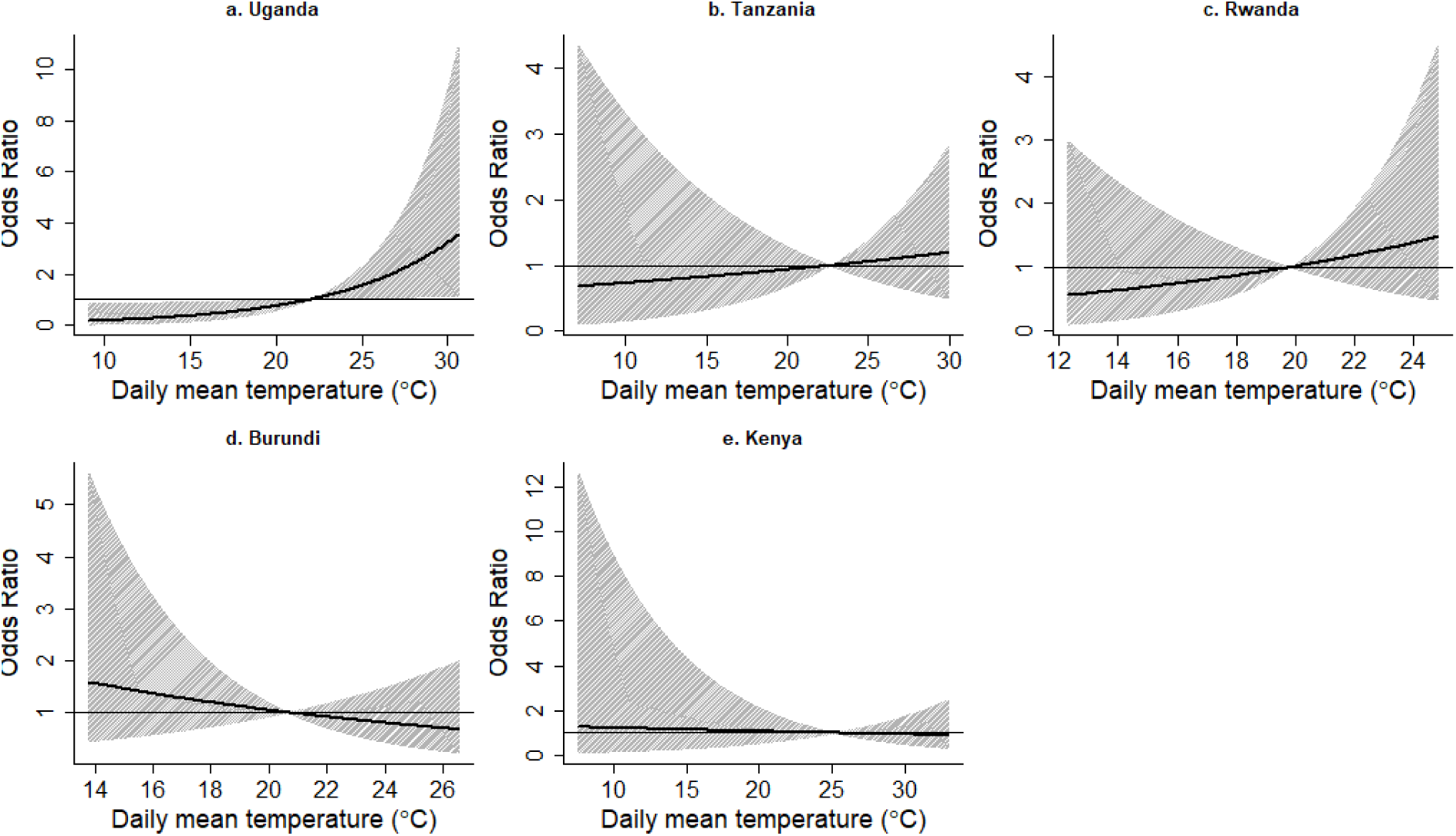
Association between the median daily temperature and the overall neonatal mortality.

Figure 2 shows the odds ratios (OR) of overall, very early, early and late neonatal mortality with exposure to the 95^th^ and 5^th^ (versus the 50^th^) temperature percentiles. Overall, there was moderate between-country heterogeneity for the effect of temperature on neonatal mortality with an overall trend towards increased mortality with exposure to high versus moderate temperatures (95^th^ vs 50^th^ percentiles) and decreased mortality with exposure to low versus moderate temperatures (5^th^ vs 50^th^ percentile). This trend was particularly notable in Uganda. For other countries, effect estimates were imprecise. Detailed forest plots for each outcome are provided in the supplementary materials.

**Figure 2:**
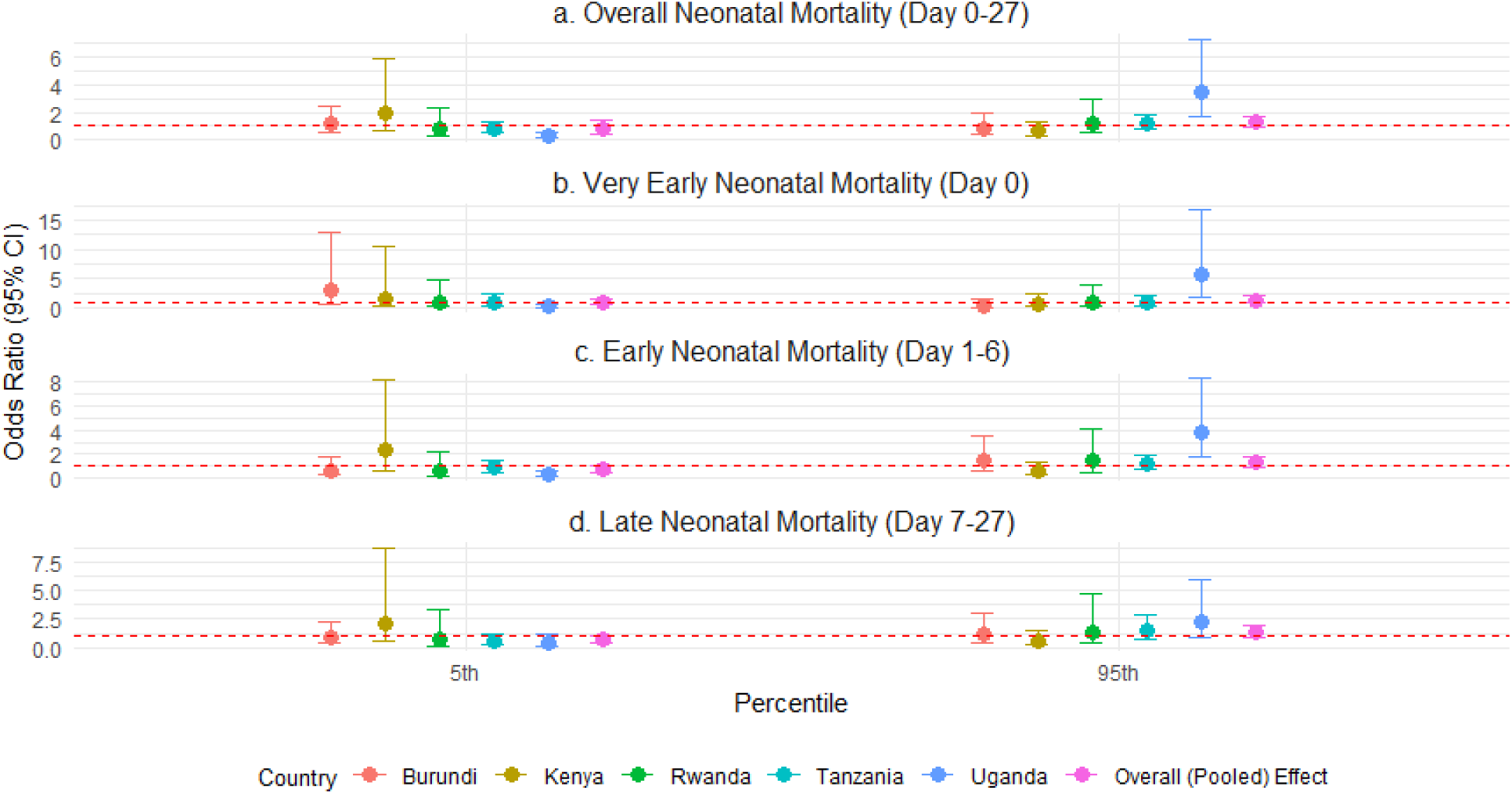
The association between the 5th and 95th percentile temperatures compared with the median temperature for the country-level overall, very early, early and late neonatal mortality.

In pooled analysis, the odds of neonatal mortality increased by 27% (OR=1.27; 95% CI: 0.95–1.70) with exposure to the 95^th^ (vs 50^th^) temperature percentile, with moderate heterogeneity (I² = 66.0%), and decreased by 22% (OR=0.78; 0.42–1.43) with exposure to the 5^th^ (vs 50^th^) percentile, also with moderate heterogeneity (I² = 64.0%). For very neonatal mortality (Figure 2b), the OR at the 95^th^ percentile was 1.23 (0.75–2.03, I² = 65.2%) and at the 5^th^ percentile was 0.78 (0.45–1.35, I² = 63.8%). For early neonatal mortality (Figure 2c), the OR at the 95^th^ percentile was 1.33 (0.96–1.84, I² = 65.6%) and at the 5th percentile was 0.69 (0.48–1.00, I² = 61.5%). The OR for late neonatal mortality (Figure 2d) was 1.29 (0.87– 1.92) at the 95th percentile, with negligible heterogeneity (I² = 4.9%), and 0.73 (0.47–1.11) at the 5th percentile, also with negligible heterogeneity (I² = 0.0%). In Uganda, a statistically significant association was observed between high environmental temperature and overall neonatal mortality (OR = 3.54; 95% CI: 1.73–7.28; Figure 2a). Similar associations were observed with very early neonatal mortality at the 5th percentile (5.65; 1.89–16.69; Figure 2b) and early neonatal mortality at the 95th percentile (3.75; 1.70–8.28; Figure 2c). Detailed forest plots are provided in the supplementary materials **ST5 – ST 12**.

Figure 3 shows the ORs of the pooled overall, very early, early and late neonatal mortality with daily mean temperatures over the study period. Overall, there was moderate between-country heterogeneity for the effect of temperature on neonatal mortality with an overall trend towards increased mortality with high temperatures and decreased mortality with low temperatures, although results were imprecise. Detailed forest plots for each outcome are provided in the supplementary materials **ST 13 – ST 14**.

**Figure 3:**
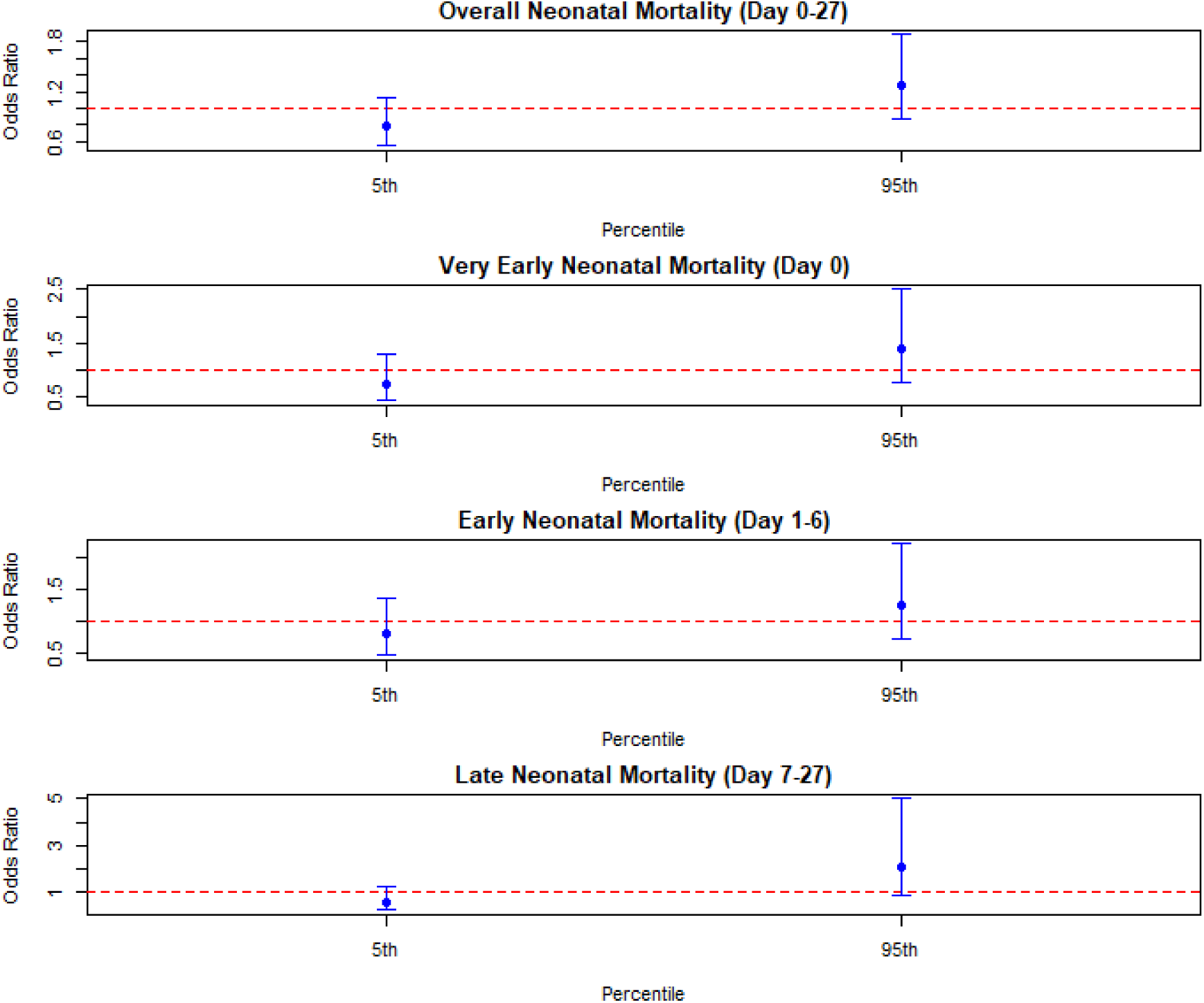
The association between the 5th and 95th percentile temperatures compared with the median temperature for the pooled overall, very early, early and late neonatal mortality.

## Subgroup analysis

The subgroup analysis results were imprecise for all the tertiles as shown in Figure 4. However, for the middle– and high-income groups, exposure to the 95^th^ and 5^th^ temperature percentiles were invariably associated with increased and decreased odds of mortality, respectively, although effect estimates results were imprecise due to small sample size. The effects of temperature were much more attenuated for both the 95^th^ and 5^th^ percentiles among the low-income group.

**Figure 4:**
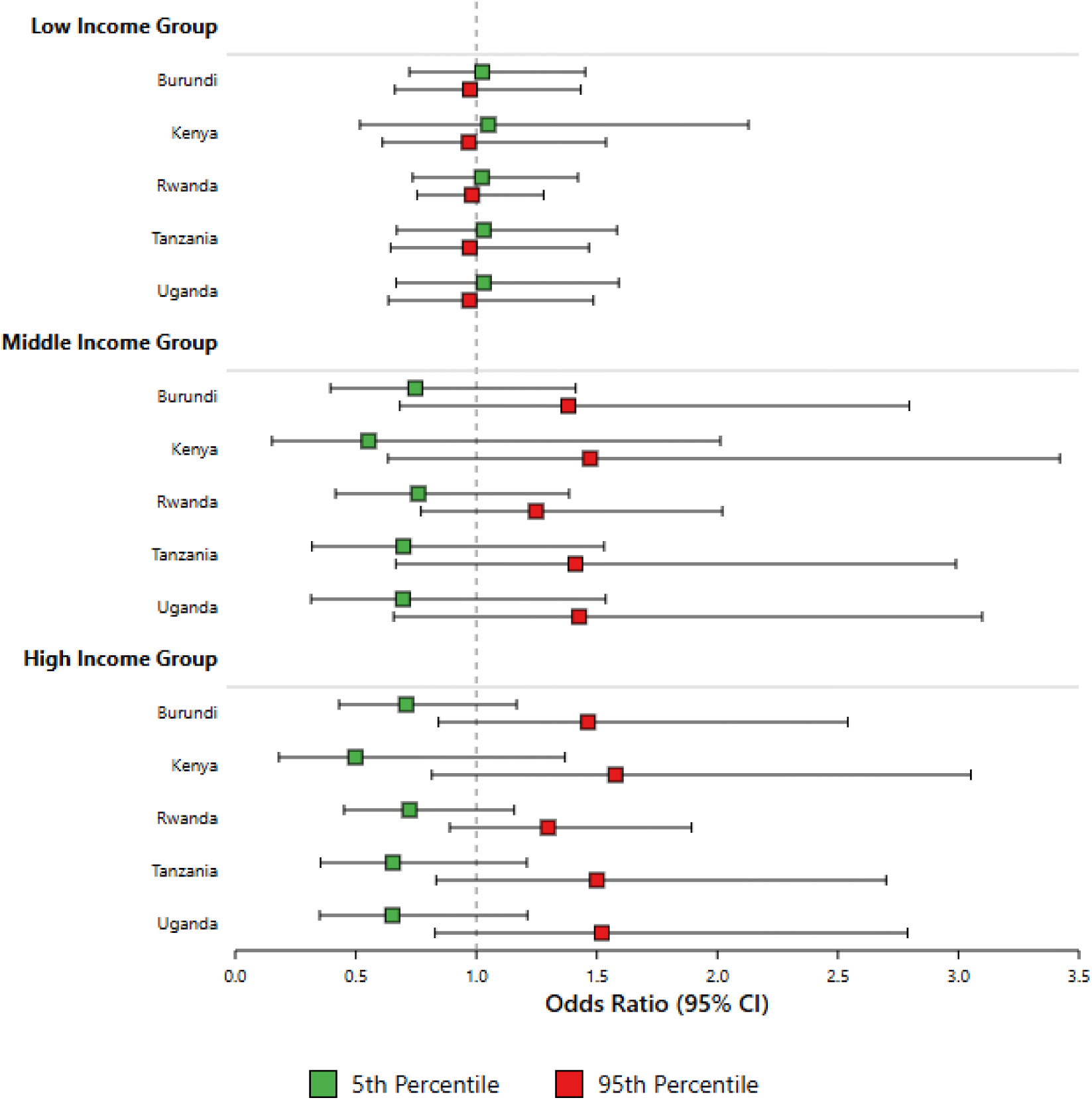
Sub-group analysis by income group.

## Discussion

In this analysis, we found that overall, higher (95^th^ percentile) temperatures were associated with increased odds of mortality while lower (5^th^ percentile) temperatures were associated with decreased of mortality for the overall, very early (day 0), early (day 1-6), and late (day 7-27) neonatal mortality. However, strong effects were only observed in Uganda while results were imprecise for the other four countries. In Uganda, higher temperatures were associated with increased odds of overall, very early and early neonatal mortality while lower temperatures associated with decreased mortality in these groups. Imprecise results were observed for the late neonatal mortality. There was no evidence of effect modification by the wealth groups.

The increased and decreased odds of mortality associated with higher and lower temperatures respectively for the overall, very early and early neonatal mortality in Uganda is consistent with other studies that have explored the relationship between temperature and neonatal mortality[5, 14, 15, 20]. The pathophysiological mechanisms through which higher temperatures lead to increased risk of neonatal mortality are still unknown. Suboptimal temperatures, both hot and cold, can lead to neonatal mortality through a variety of direct and indirect mechanisms. Direct mechanisms primarily involve the neonate’s immature thermoregulatory system[6, 46–48]. Neonates, especially preterm and low birth weight infants, have a narrower optimal body temperature range than adults[5, 6]. Their higher metabolic rate and lower sweating rate limit their ability to dissipate heat effectively[5], making them vulnerable to hyperthermia. Conversely, their high surface-area-to-mass ratio increases the risk of rapid heat loss and hypothermia[5, 14].

High ambient temperatures have been linked to preterm birth, placental abruption, and gestational hypertension, all of which increase the risk of overall neonatal mortality[3, 5, 46, 49]. However, there may be different medical causes according to the age of the newborns, [5]. Extreme temperature exposure has been linked with prematurity, low birth weight, maternal dehydration, and other childbirth complications, which increases risk of very early neonatal mortality[3, 6, 14, 19, 20]. On the other hand, late neonatal mortality may be due to severe infections such as sepsis and pneumonia, which are often associated with hypothermia[50–52].

Health systems in low-resource settings may be ill-equipped to manage the increased burden of neonatal complications arising from temperature extremes. Lack of access to essential equipment like incubators and infant warmers, as well as unreliable power supplies, can exacerbate the risks of both hypothermia and hyperthermia[5]. Beyond the immediate physiological impacts, sub-optimal temperatures can indirectly influence neonatal mortality by exacerbating existing environmental and socioeconomic vulnerabilities. Heat can negatively impact healthcare system responsiveness by contributing to healthcare worker exhaustion and disrupting agricultural production, ultimately affecting nutrition and access to care[46]. Moreover, heat waves can directly impede access to essential healthcare services, leading to delays in care-seeking and potentially fatal consequences for vulnerable neonates[53]. The broader impacts of climate change, including extreme weather events such as droughts and floods, further compound these risks. These events can damage critical infrastructure, displace populations, and disrupt food supplies, leading to increased rates of malnutrition and infectious diseases in neonates[53].

In our analysis, we noted between-country heterogeneity on the effect of temperature on neonatal mortality. These results are similar to findings by Brimicombe et al. (2024) in which no statistically significant effect of the 95^th^ percentile was detected compared with the median heat exposure in the East Africa region (OR = 1.02, 95% CI: 0.96-1.08)[20].

However, their study pooled the East African region countries and therefore, unlike this analysis, between-country results are not teased out. These findings are also similar to findings by Dimitrova et al. (2024) in as while they showed that 4.3% of all neonatal mortalities across 29 LMICs were associated non-optimal temperatures (hot or cold), the effects across Burundi, Rwanda, Tanzania and Uganda were not statistically significant[5]. Furthermore, a scoping review examining the effect of elevated ambient temperature on maternal, foetal and neonatal outcomes found mixed effects, with increased, decreased or no risk in neonatal mortality[15]. Similar findings were reported in the systematic review by Lakhoo et al. (2022) which reported mixed findings on the effect of heat on neonatal mortality across different regions and climate zones[14].

The apparent insufficient evidence of the association between non-optimal temperatures and neonatal mortality in the East Africa region could be explained by several factors. First, the 5^th^ and 95^th^ temperature percentiles, chosen in our study as the cold and hot temperatures respectively, ranged from 16.1 – 17.6°C and 22.8 – 30.4°C respectively, did not differ considerably from the median temperature, our centering value, which ranged from 19.8°C–25.2 °C. Generally, the countries had narrow temperature ranges. Secondly, the number of cases (mortalities) in our data set was small which may have led to increased sampling errors and wider confidence intervals[54]. Moreover, the number of cases at the 95^th^ and 5^th^ percentiles were even lower. Thirdly, there could be factors other than temperature, such as preterm births and birth asphyxia, responsible for neonatal mortality in the East African region[20], although these are also linked with extreme temperatures. Finally, health care practices such as care for the newborn which empathise thermal care including Kangaroo Mother Care, especially for the low– and preterm births may attenuate the effect of ambient temperature on neonatal mortality in these settings[55, 56].

Our analysis draws on a number of strengths. First, by employing a time-stratified case-crossover design, we controlled for time-invariant confounders, minimising the risk of confounding in our results. Secondly, we analysed data per country, rather than at regional or global level, which often masks the differential effects of temperature at the regional and/or global level, including confounders that may vary between countries. While this does not completely disaggregate data by fine temperature zones, it provides much more detailed country-specific and between-country results. Finally, to the best of our knowledge, this is the first study that has explored the effects of temperature on neonatal mortality at different time periods, namely very early, early and late neonatal mortality.

## Limitations

However, our study suffers from the following limitations. Firstly, given the relative rarity of the outcome, the sample size, as reported above was very small which may have affected the confidence intervals. Secondly, in the DHS analysis, the GPS coordinates are linked to the PSU. No information about the place of mortality or where the child was is provided[23]. Thus, it is possible that the child could have died in an area with a different temperature compared with what we used in the analysis. However, we do anticipate the mean daily temperature not to vary greatly between the DHS strata[25]. Finally, our analysis may suffer from recall bias since the information about births and mortality is collected for the five years preceding the survey. This means that we may have included stillbirths as neonatal deaths and/or misclassified neonatal deaths by the period of death. We tried to minimise this by focusing on the most recent survey.

## Conclusion and recommendation

While our overall results show heterogeneity and uncertainty, we note that higher temperatures were generally associated with increased odds of mortality compared with lower temperatures. This may be due to the fact that the impact of cold on neonates is well established and health providers and families are well trained in reducing cold stress in infants, taking extra care during low temperature days. There is therefore a need to incorporate heat mitigation strategies into the care for newborn policies and practices, especially for the very early and early neonatal periods. There is also a need to develop and implement country-specific and age-specific guidelines for thermal management of neonates, especially in Uganda.

In conclusion, temperature-related neonatal mortality risk varies across East African countries, with only Uganda showing significant associations with high and low temperatures. The effects of temperature on neonatal mortality may be more pronounced in the first 6 days of life, perhaps linked to childbirth complications, which may be a sequelae of suboptimal temperature exposure during pregnancy. The findings also suggest that other factors may play a more significant role in neonatal mortality in East Africa. There is a need for further research to understand policies, practices and mechanisms underlying the relationship between temperature and neonatal mortality in Uganda. There is also a need to explore the effects of temperature on neonatal mortality by the different climate/temperature zones.

## Contributors

PL is the guarantor. PL and VF conceived the project. PL, CP, and CB provided the analysis plan. SK, DJ, HB, and VF reviewed and approved the final analysis plan. PL analysed the data and wrote the first draft. All remaining authors edited the draft and provided critical inputs. All authors have approved and contributed to the final manuscript submitted. PL had the final responsibility for the decision to submit for publication.

## Patient consent for publication

Not required.

## Data availability statement

All the data generated or analysed during this study can be obtained with written permission from the DHS Program (https://dhsprogram.com).

## Acknowledgements

HIGH Horizons has received funding from the European Union’s Horizon Framework Programme under Grant Agreement No. 101057843. Project partner LSHTM is funded by UKRI Innovate UK reference number 10038478. We thank the HIGH Horizons study group, namely Stanley Luchters, Gloria Maimela, Celeste Madondo, Shobna Sawry, Mags Beksinska, Lebohang Radebe, Ijeoma Solarin, Pascalia Munyewende, Chuansi Gao, Jakob Eggeling, Gunter Alce, Clara Heil, Nathalie Roos, Olof Stephansson, Claudia Hanson, Veronika Tirado, Anayda Portela, Jorn Toftum, Sohail Baloch, Jetina Tsvaki, Thabani Moronzie, Fortunate Machingura, Concilia Mutasa, Brian Mgondisi Sibanda, Tariro Chinozvina, Elizabeth Dangaiso, Jasper Maguma, Bongani Mutimutema, Veronique Filippi, Giulia Greco, Nasser Fardousi, Isabelle Lange, Giorgia Gon, Jo Borghi, Christo Hadjichristodoulou, Barbara Mouchtouri, Elina Kostara, Maria Kyritsi, Michalis Koureas, Fani Kalala, Chara Bogogiannidou, Ioanna Voulgaridi, Boris Kingma, Koen van der Sanden, Federica Noble, and Massimo Stafoggia.

## Supplementary materials

**ST 1:**
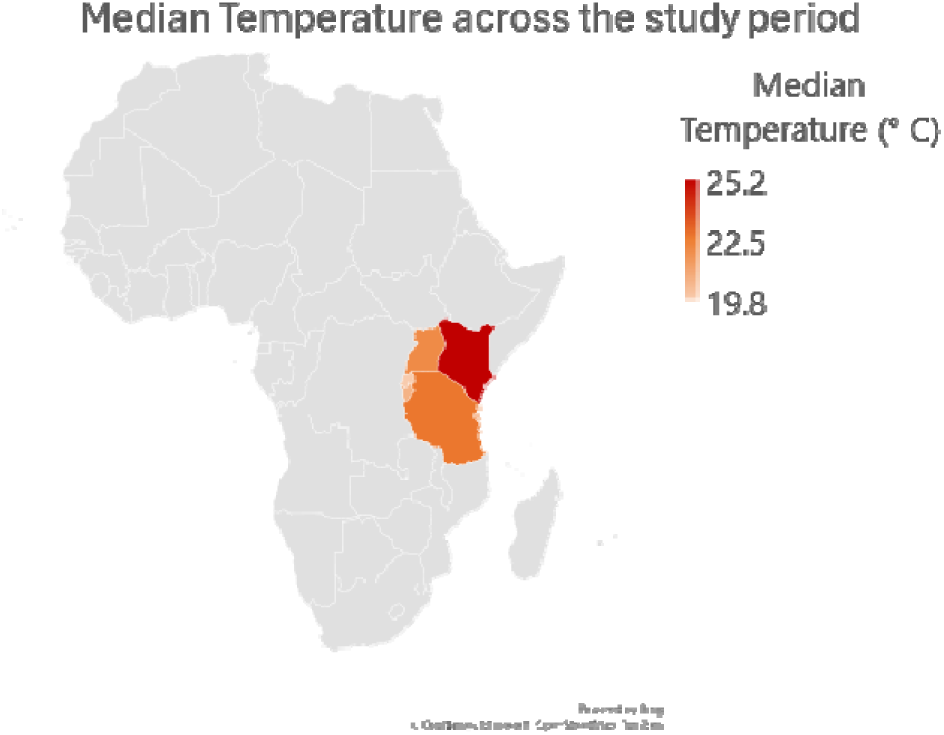
Map showing median temperature over the East African region.

**ST 2:**
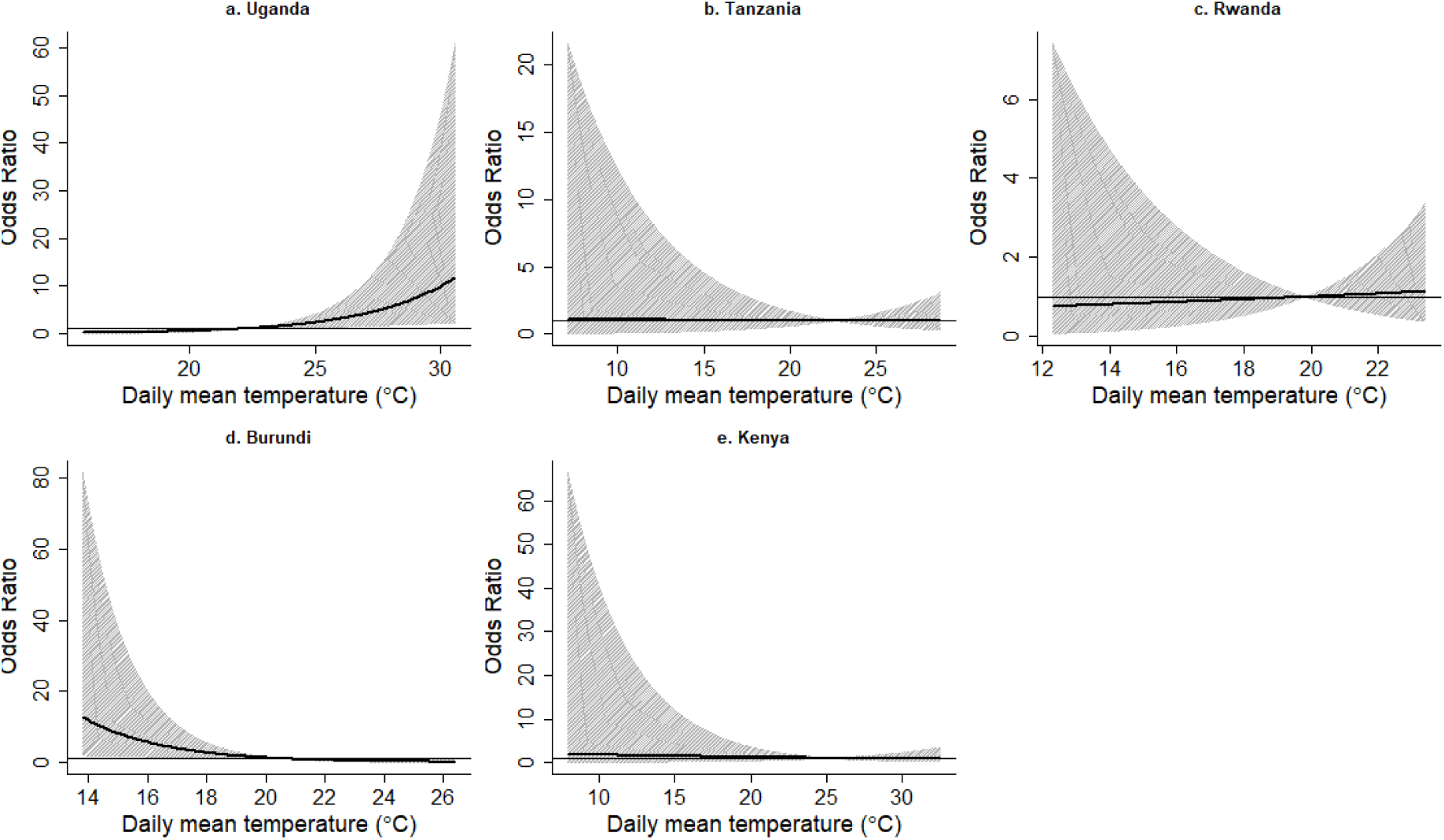
Association between the median daily temperature and very early (day 0) neonatal mortality.

**ST 3:**
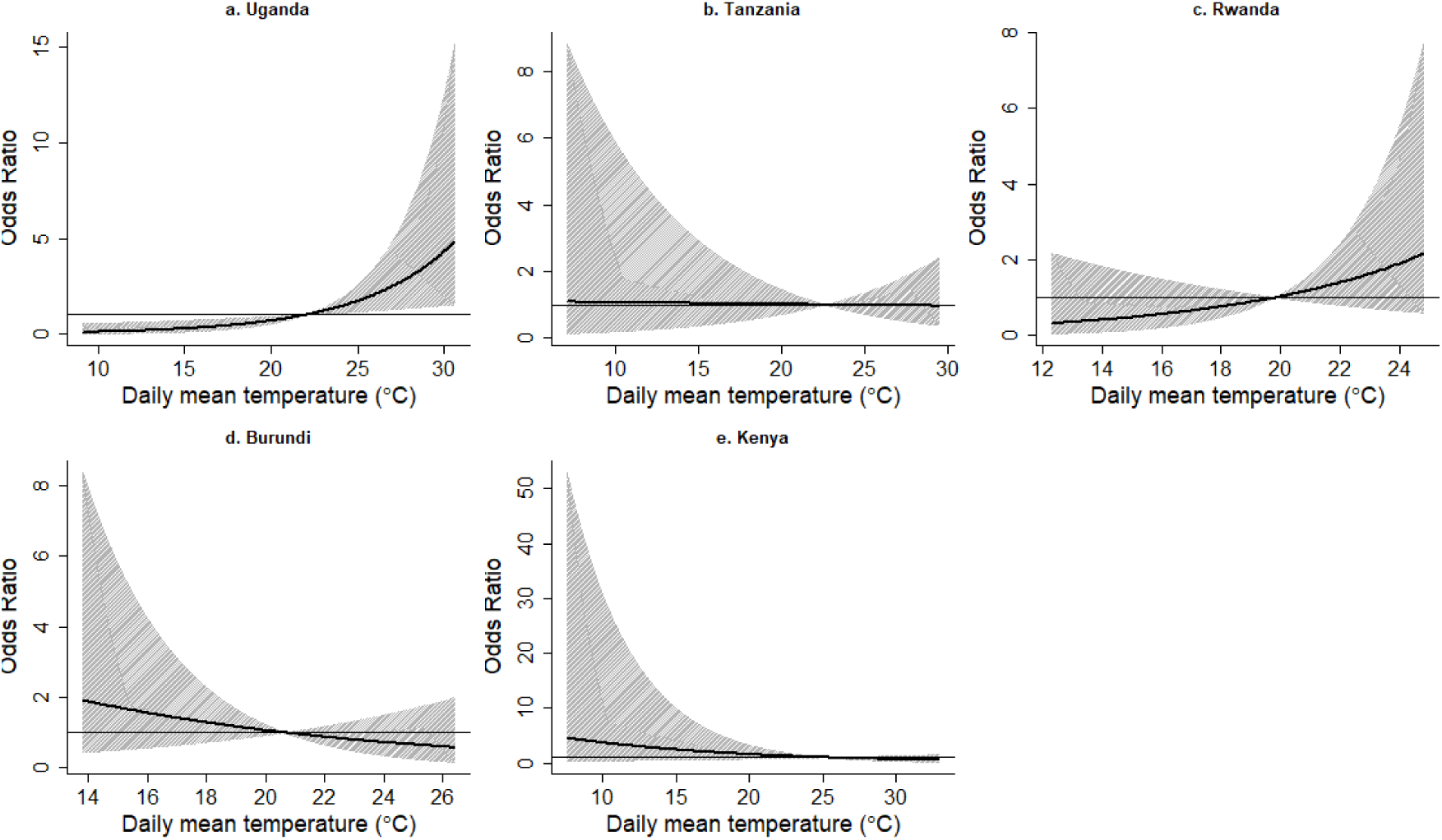
Association between the median daily temperature and early (day 1-6) neonatal mortality.

**ST 4:**
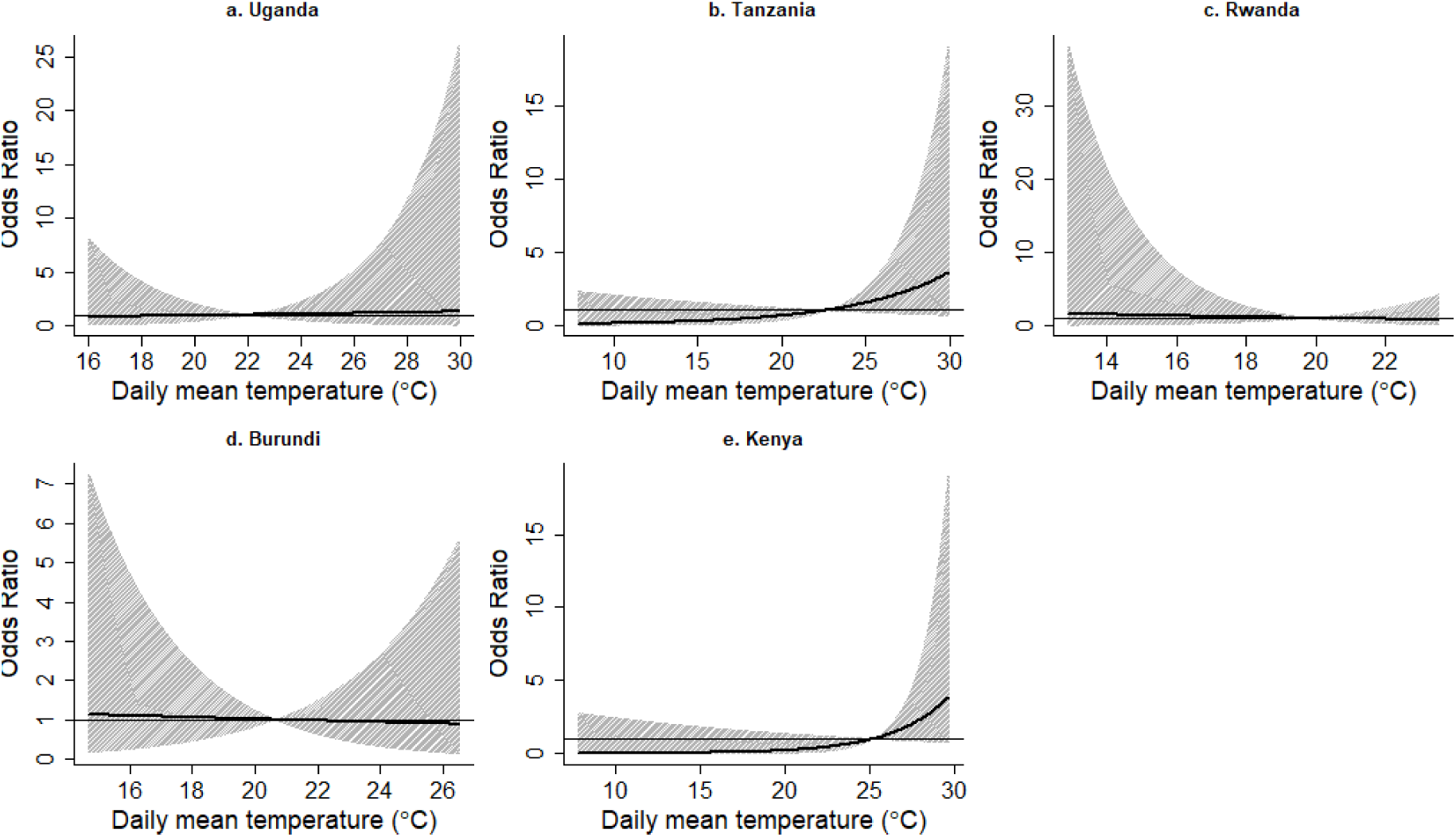
Association between the median daily temperature and late (day 7-27) neonatal mortality.

**ST 5:**
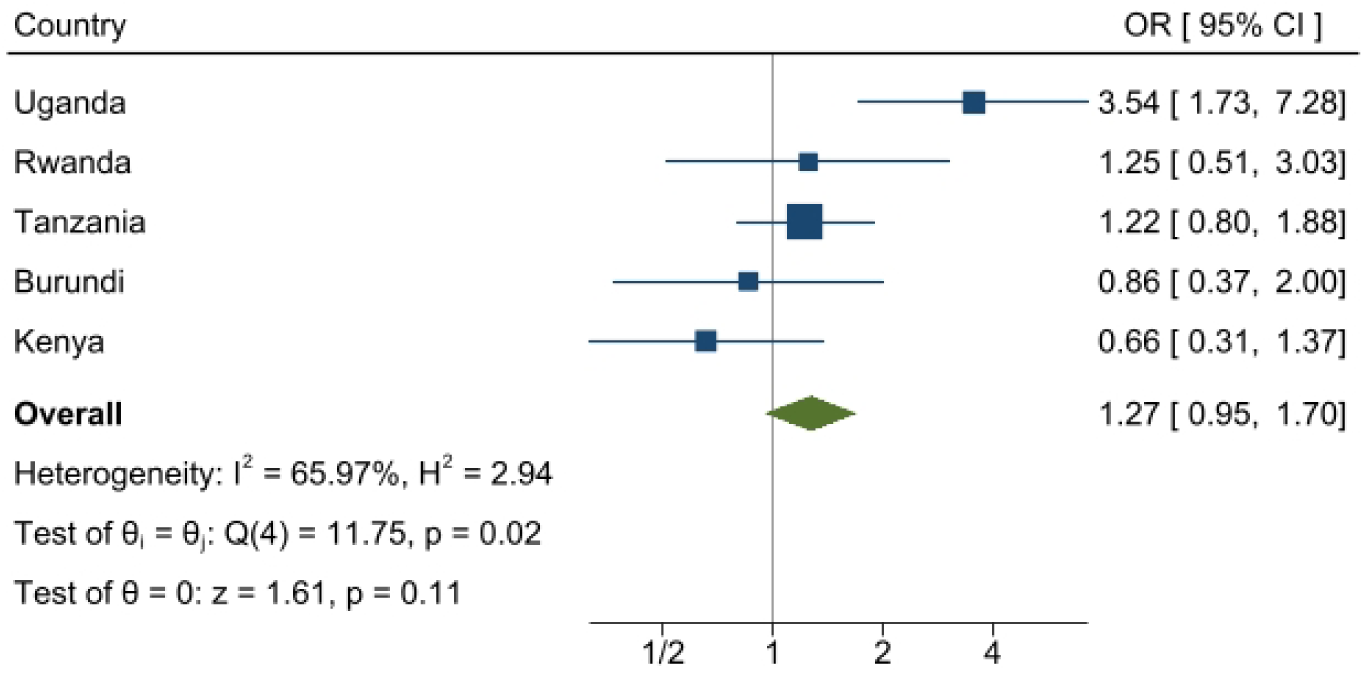
The overall neonatal mortality: The forest plot for 95th percentile vs median cumulative effects.

**ST 6:**
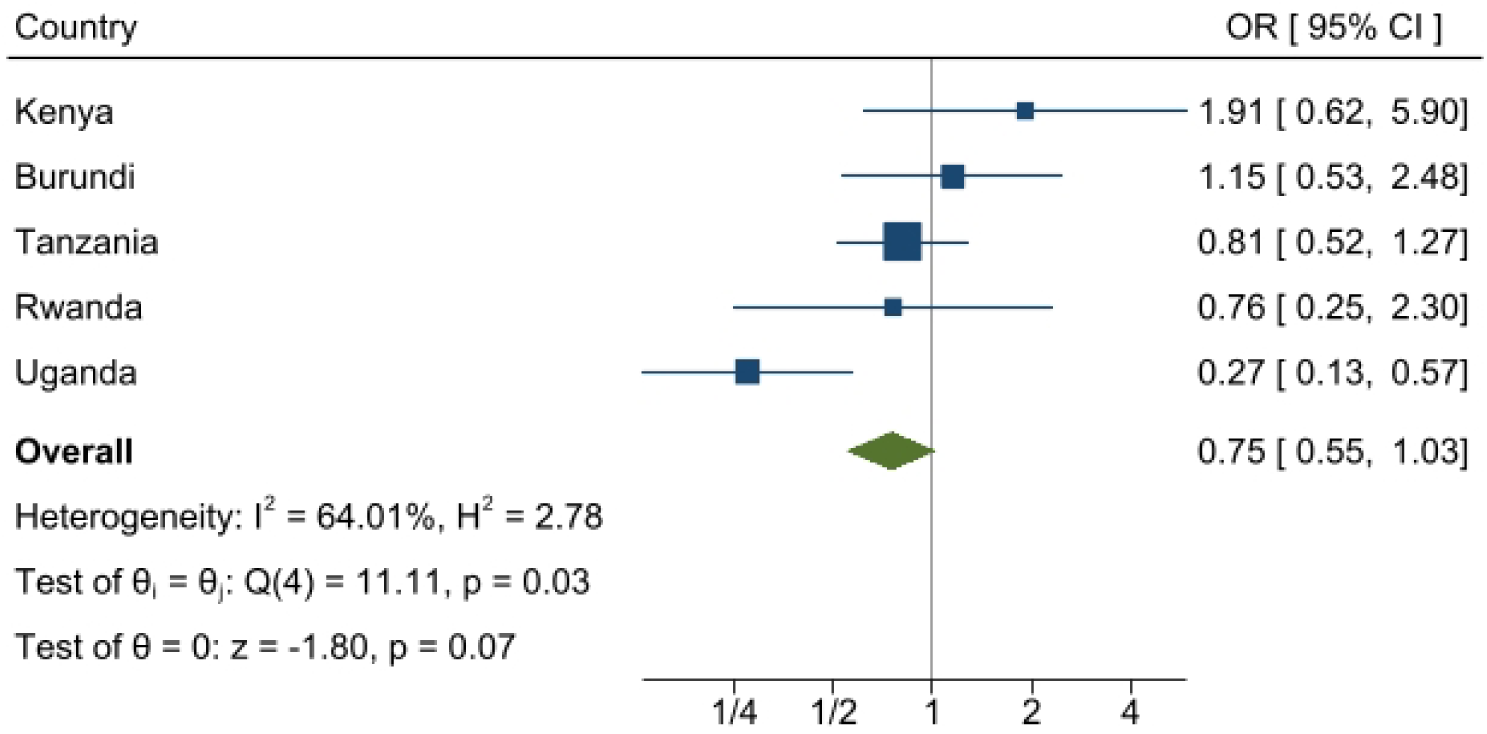
The overall neonatal mortality: The forest plot for 5th percentile vs the median.

**ST 7:**
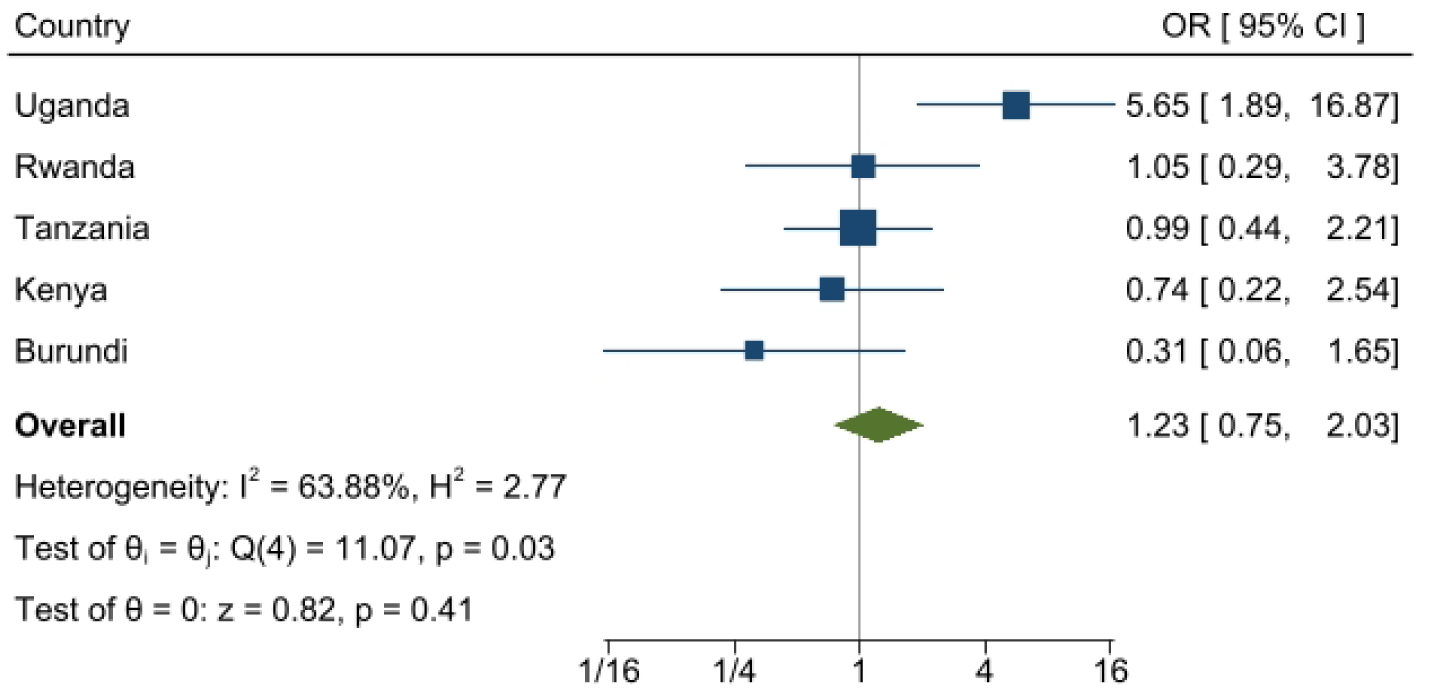
Very early neonatal mortality: 95th percentile vs the median.

**ST 8:**
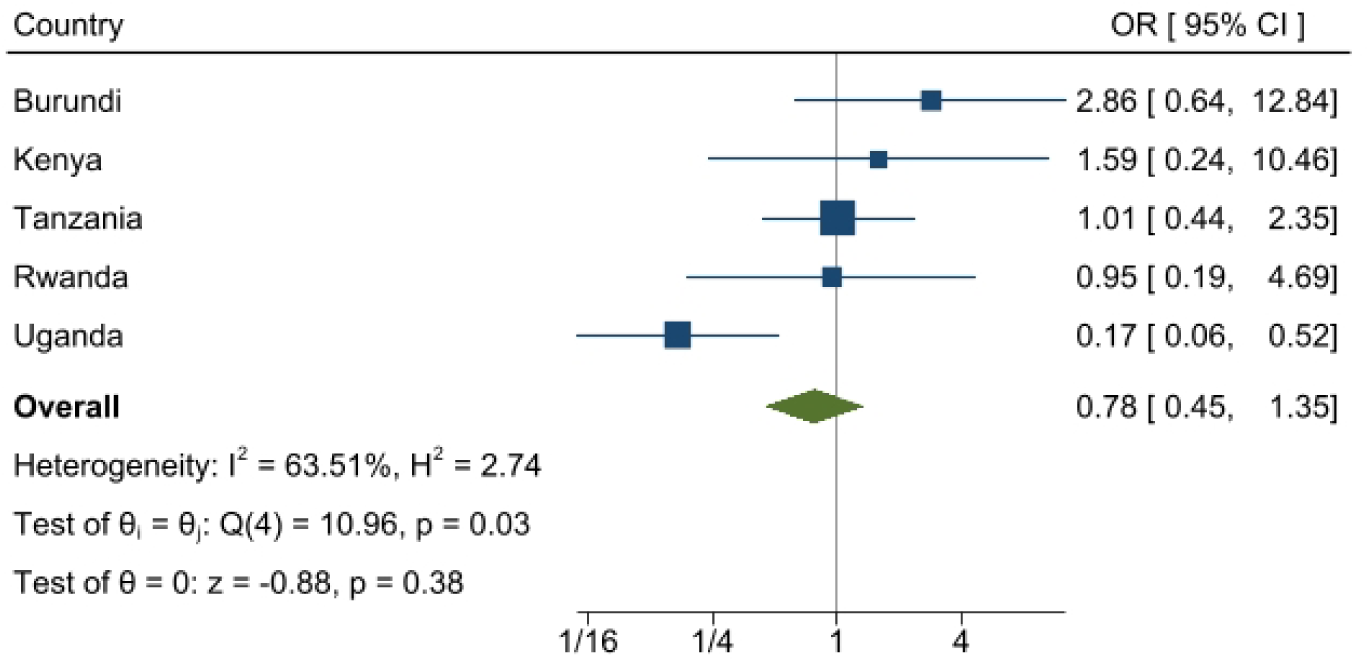
Very early neonatal mortality: 5th percentile vs the median.

**ST 9:**
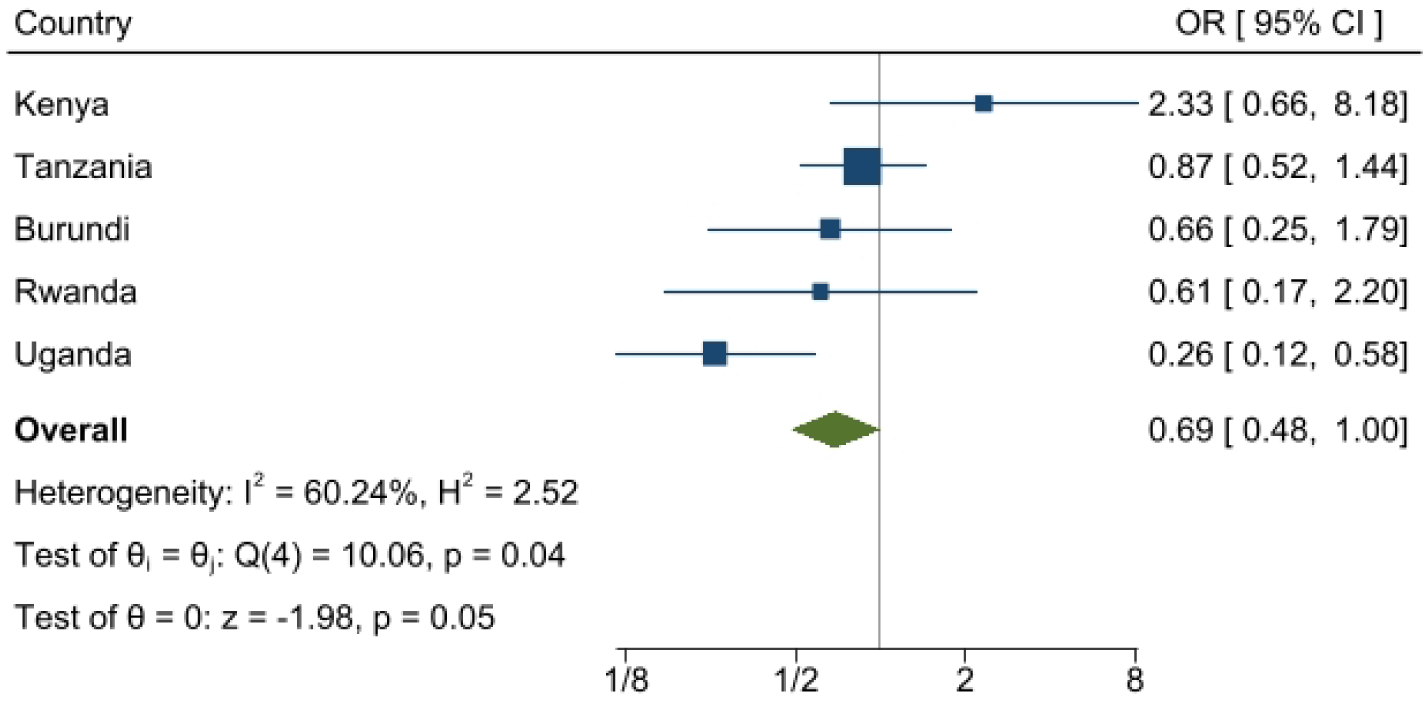
Early neonatal mortality: 95th percentile vs the median.

**ST 10:**
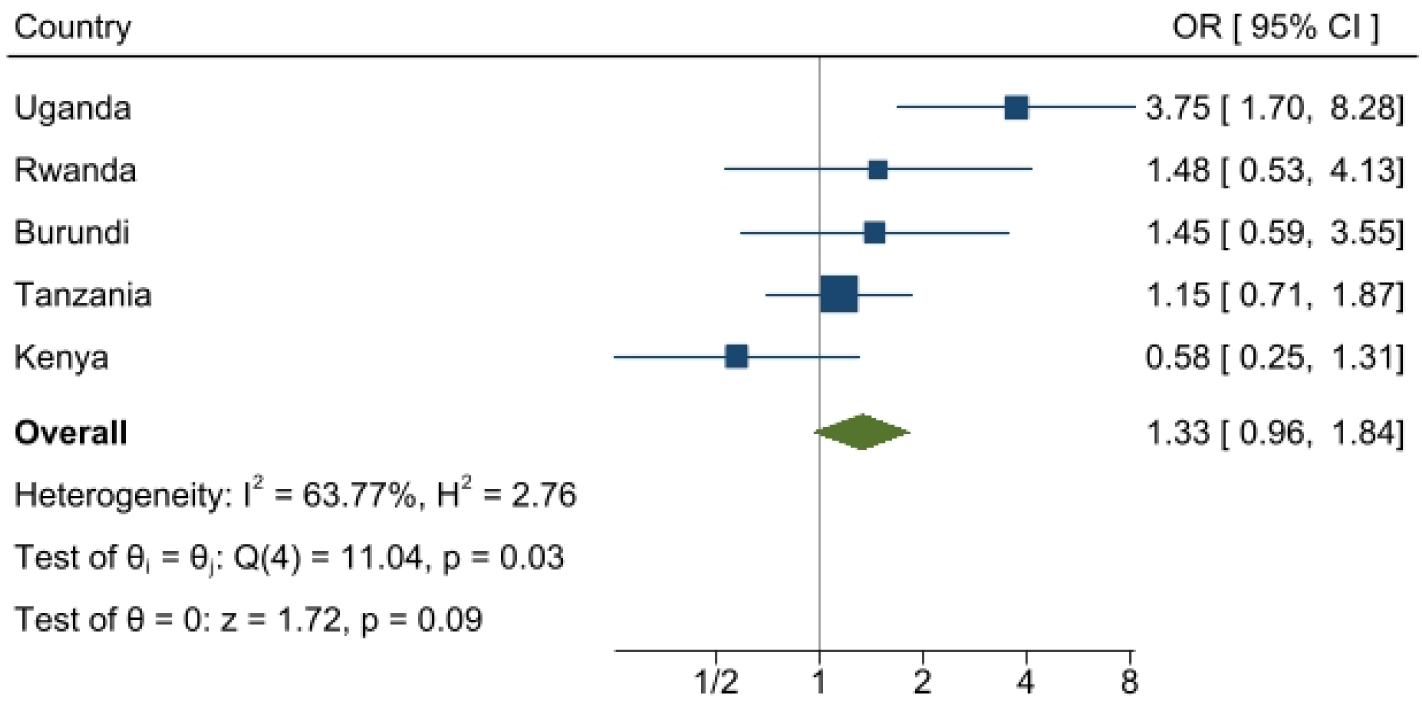
Early neonatal mortality: 5th percentile vs the median.

**ST 11:**
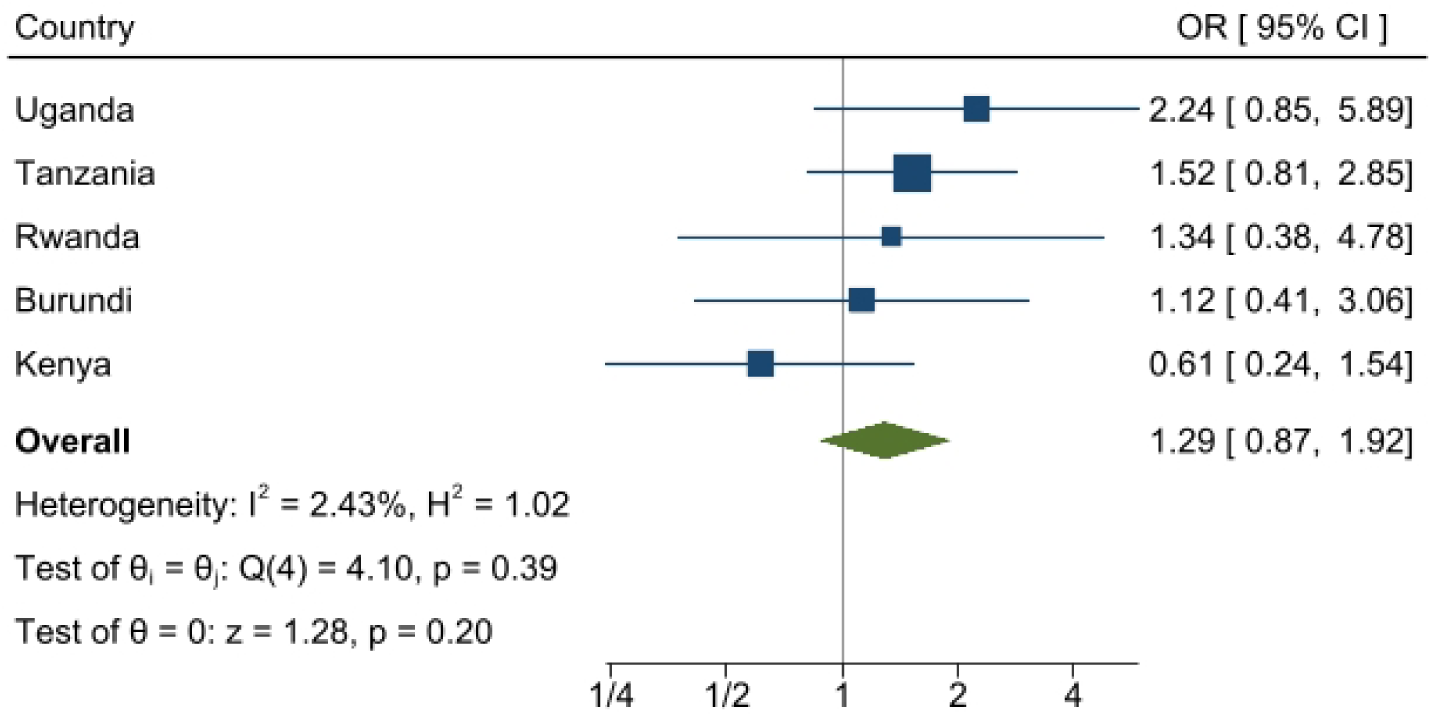
Late neonatal mortality: 95th percentile vs the median.

**ST 12:**
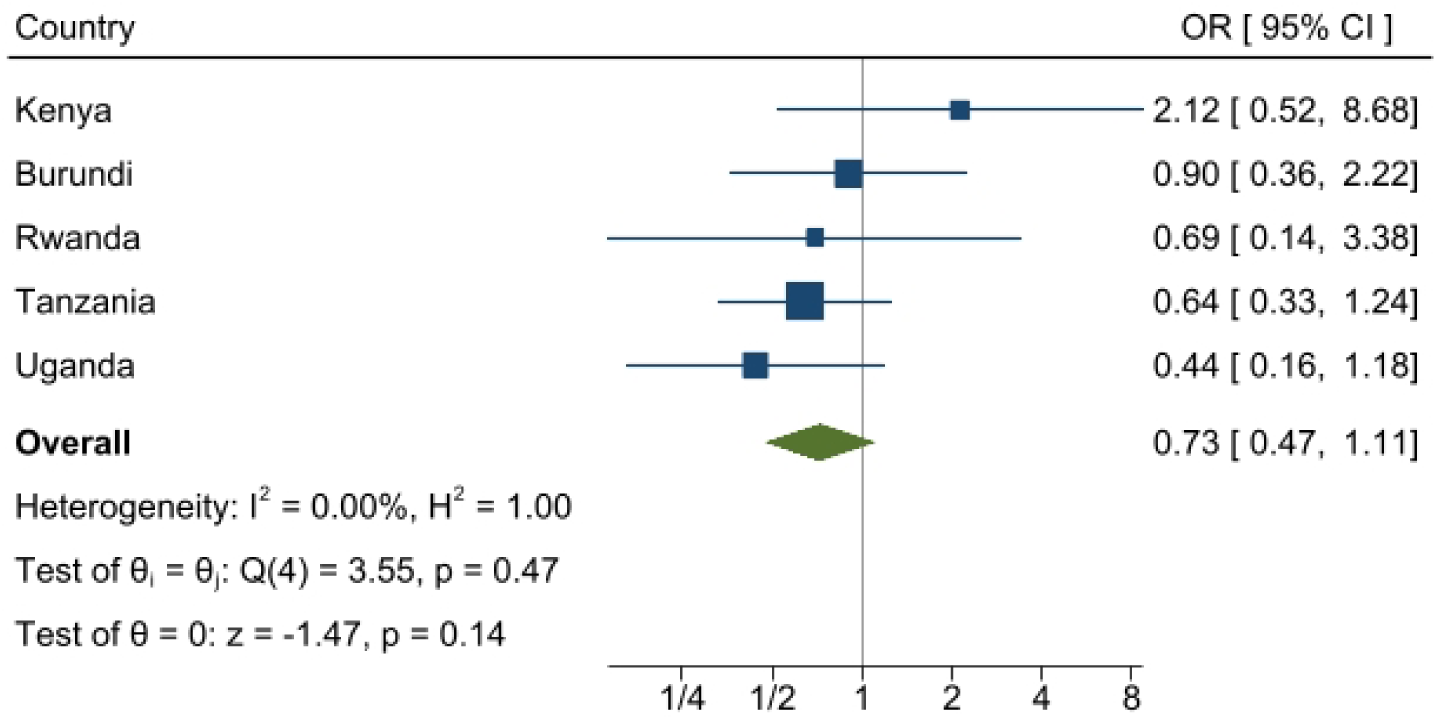
Late neonatal mortality: 5th percentile vs the median.

**ST 13:**
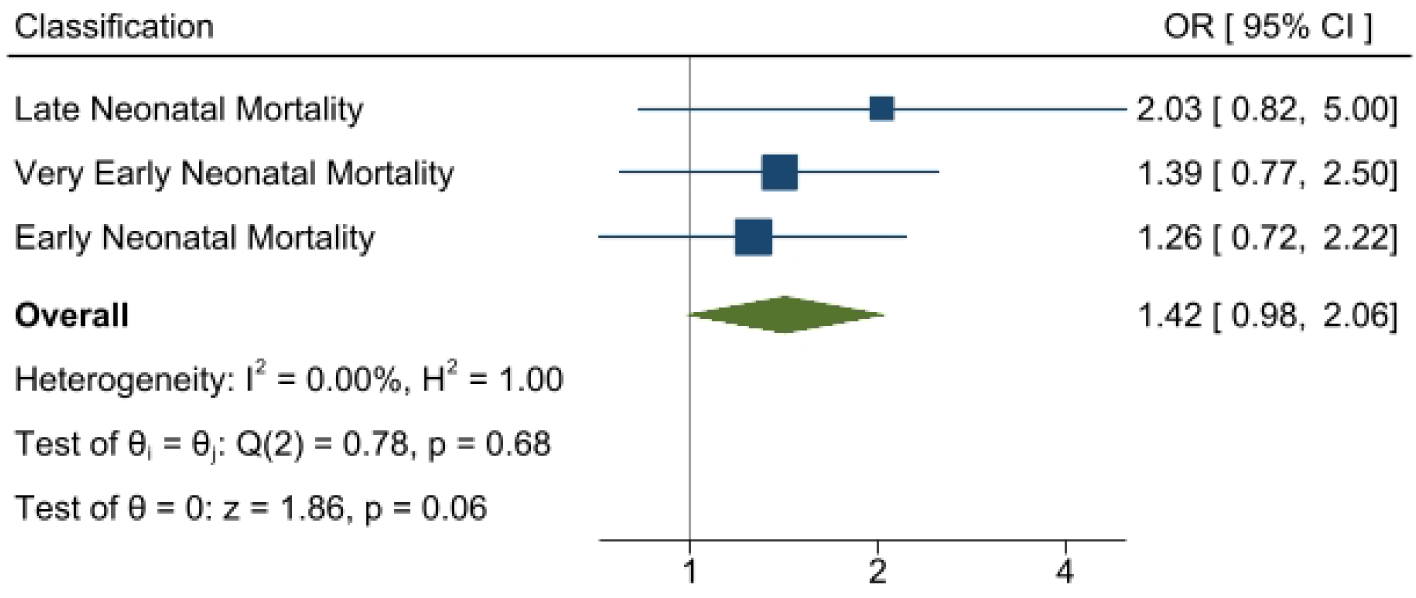
Overall effect for very early, early and late neonatal mortality, 95th percentile vs the median.

**ST 14:**
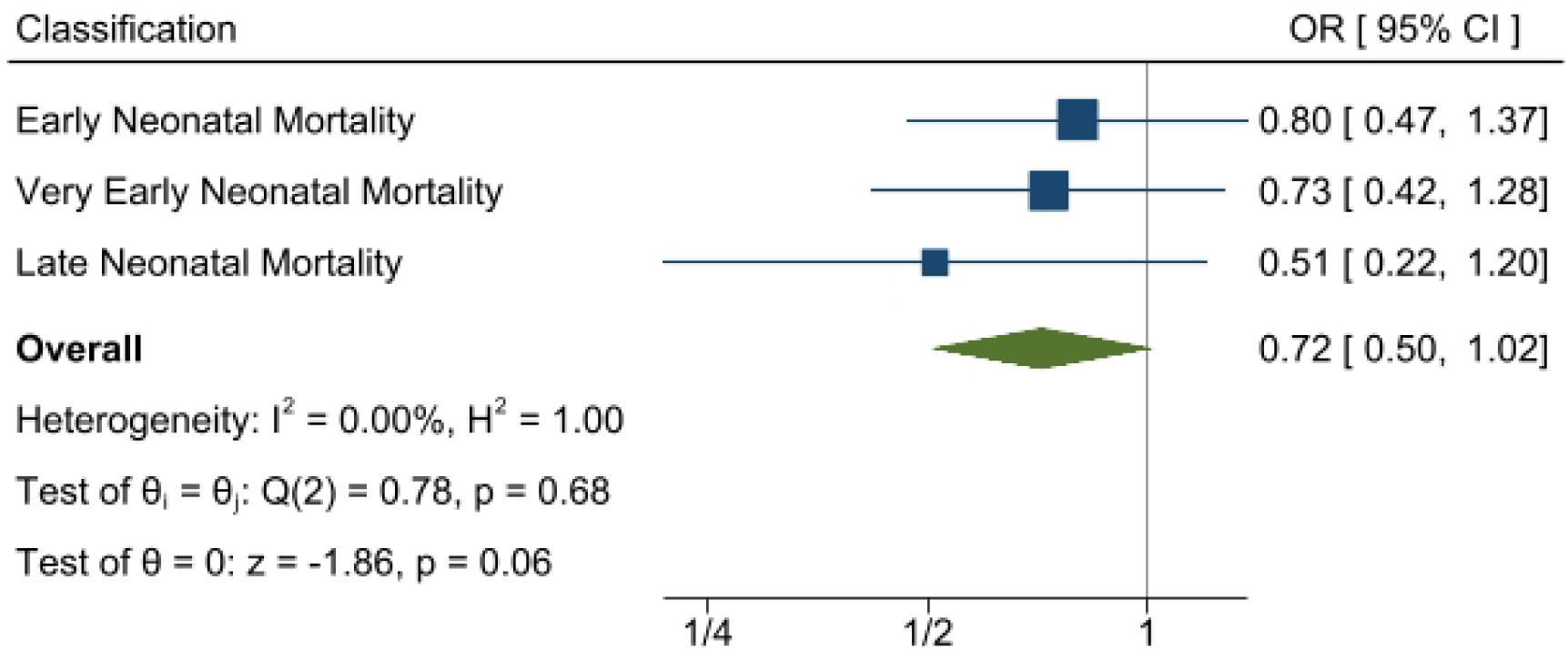
Overall effect for very early, early and late neonatal mortality, 5th percentile vs the median.

